# Three Months of Partnered Dance Aerobic Exercise Can Reduce OFF-time and Enhance Quality of Life in Older Adults with Parkinson’s Disease

**DOI:** 10.1101/2025.07.17.25331727

**Authors:** Caroline L. Kim, David Morton, J. Lucas McKay, Joe R. Nocera, Daniel Huddleston, Forouzan Rafie, Madeleine E. Hackney

## Abstract

**Introduction:** Parkinson’s Disease (PD) is commonly treated with the dopamine precursor, levodopa, which is used in antiparkinsonian therapy. After several years of taking this medication, many individuals with PD experience medication-related motor fluctuations (MRMF), a.k.a., OFF-time. OFF-time is one of the most disconcerting features of PD when troubling motor and non-motor symptoms previously alleviated by levodopa return. Exercise, specifically dance, could beneficially address OFF-time and enhance quality of life (QOL) and independence in people with PD.

**Methods:** This assessor-blinded randomized controlled trial (1:1) [NCT04122690] included 45 participants with PD (70.1±7.27 years, 73% male, 2.30±0.61 Hoehn & Yahr stage), who were randomized into 30 hours of PDAE or WALK over 3 months. Psychosocial questionnaires, MDS-UPDRS, and 3-day OFF-state diaries were administered at baseline and three-month timepoints to measure reported OFF-time, QOL, independence, and disease severity. Within-group comparisons were analyzed using paired t-tests, and between-group comparisons were analyzed using independent t-tests and a linear mixed-effects model.

**Results:** After three months, PDAE reduced OFF-time and improved motor symptoms. Compared to WALK, PDAE also reduced OFF-time, improved motor symptoms, and enhanced the experience of daily living.

**Conclusion:** PDAE is superior to WALK and is an effective adjunctive therapy to help improve OFF-time and QOL in individuals with PD after three months. Further studies are needed to determine the relationship between improving OFF-time and quality of life.

## Introduction

Parkinson’s Disease (PD) is an intractable condition that severely impairs motor and cognitive function as well as quality of life and overall wellbeing. Neurologically, PD is characterized by a severe loss of dopaminergic neurons in the pigmented pars compacta of the substantia nigra in the midbrain [1]. After several years of dopamine replacement, levodopa, treatment, up to 75% of PD patients experience medication-related motor fluctuations, which is also known as “OFF-time”: the wearing-off of levodopa and reoccurrence of troubling motor and non-motor symptoms [2]. OFF-time is one of the most disconcerting features of PD because of its impact on the individual’s quality of life (QOL) [3]

Common functional problems of people with PD affect motor, cognitive, and psychosocial domains. Motor issues include akinesia (difficulty initiating movements), bradykinesia (slow movements), rigidity, and tremor. Cognitive problems are one of the more significant nonmotor symptoms of PD with insidious onset [4]. Cognitive deficits in PD significantly impact executive function, speech dysfunction, visual-spatial ability, and memory impairment [5]. Other nonmotor symptoms of PD include depression, dementia, and psychosis, and have a substantial impact on QOL. PD also impacts independence and the ability to carry out activities of daily living (ADL) and psychosocially may lead to depression, anxiety, and apathy, impacting the individual’s QOL and independence [6].

Currently, levodopa, a dopaminergic precursor, is the standard pharmaceutical treatment for PD and effectively improves motor symptoms in many patients within 15 to 30 minutes. Despite its ease and speed of acting within the brain, levodopa is not a perfect PD treatment. The prevalence of OFF-time is concerning due to its adverse impact on independence and QOL and is one of the most troublesome symptoms frequently reported by patients [3]. OFF-time negatively impacts non-motor and motor symptoms including mobility, activities of daily living, emotional well-being, cognition, communication, and bodily discomfort. The negative impact of OFF-time on mobility is described as a significant component of experiencing OFF-time [7].

Levodopa is commonly taken with aromatic L-amino acid decarboxylase (AADC) inhibitors like carbidopa or benserazide and catechol-O-methyltransferase (COMT) inhibitors to improve bioavailability and reduce peripheral side effects like cardiac arrhythmias, hypotension, nausea, and vomiting [8]. Despite the multitude of pharmacological options, there remain issues with each one, and patients still experience considerable OFF-time, which burdens patients’ and families’ QOL [9]. Therefore, alternative, or adjunctive therapies are necessary to help address OFF-time in individuals with Parkinson’s Disease.

Exercise has many benefits for patients with PD. It is increasingly promoted as helpful for motor and cognitive symptom relief and may be neuroprotective. Exercise also grossly affects organ tissue and function, which may influence the pharmacokinetics of drugs [10]. Dance has been shown to be a promising form of exercise for older adults, including those with PD. In general, dance has been proven to be beneficial for individuals with PD, especially for improving QOL and motor impairment, specifically balance, gait, and motor symptom severity [39]. An adapted form of the Argentine tango (adapted tango) has also proven to be beneficial for adults with PD, improving mobility and decreasing gait deficits while also improving spatial cognition [11].

In the current study, we aim to investigate the effects of three months of cognitively engaging social dance, Partnered Dance Aerobic Exercise (PDAE) when compared to a walking aerobic exercise (WALK) on the impact of OFF-time and perceived QOL and independence factors in older adults with PD. We look to determine if a three-month program of aerobic exercise in general affects OFF-time and QOL and independence factors, or if PDAE is a superior form of aerobic exercise to WALK. OFF-time symptoms and their impact on the patient’s QOL and independence factors are often associated. However, few studies examine how exercise-based interventions, specifically social dance, impact OFF-time symptoms and, consequently, QOL and independence factors. Improving OFF-time symptoms in PD patients could be a valuable key to improving their QOL and independence.

## Methods

This study was approved by the Institutional Review Board of Emory University and the Review Committee for the Atlanta Veterans Affairs Medical Center (VAMC). Participants followed the PAIRED protocol, which is a parallel, assessor-blinded RCT (1:1) that has been registered on clinicaltrials.gov (NCT04122690), and has previously been published (Hackney et al., 2020). Figure 1 describes participation throughout the study. The Consolidated Standards of Reporting Trials (CONSORT) were followed to ensure the quality of reporting.

**Figure 1.**
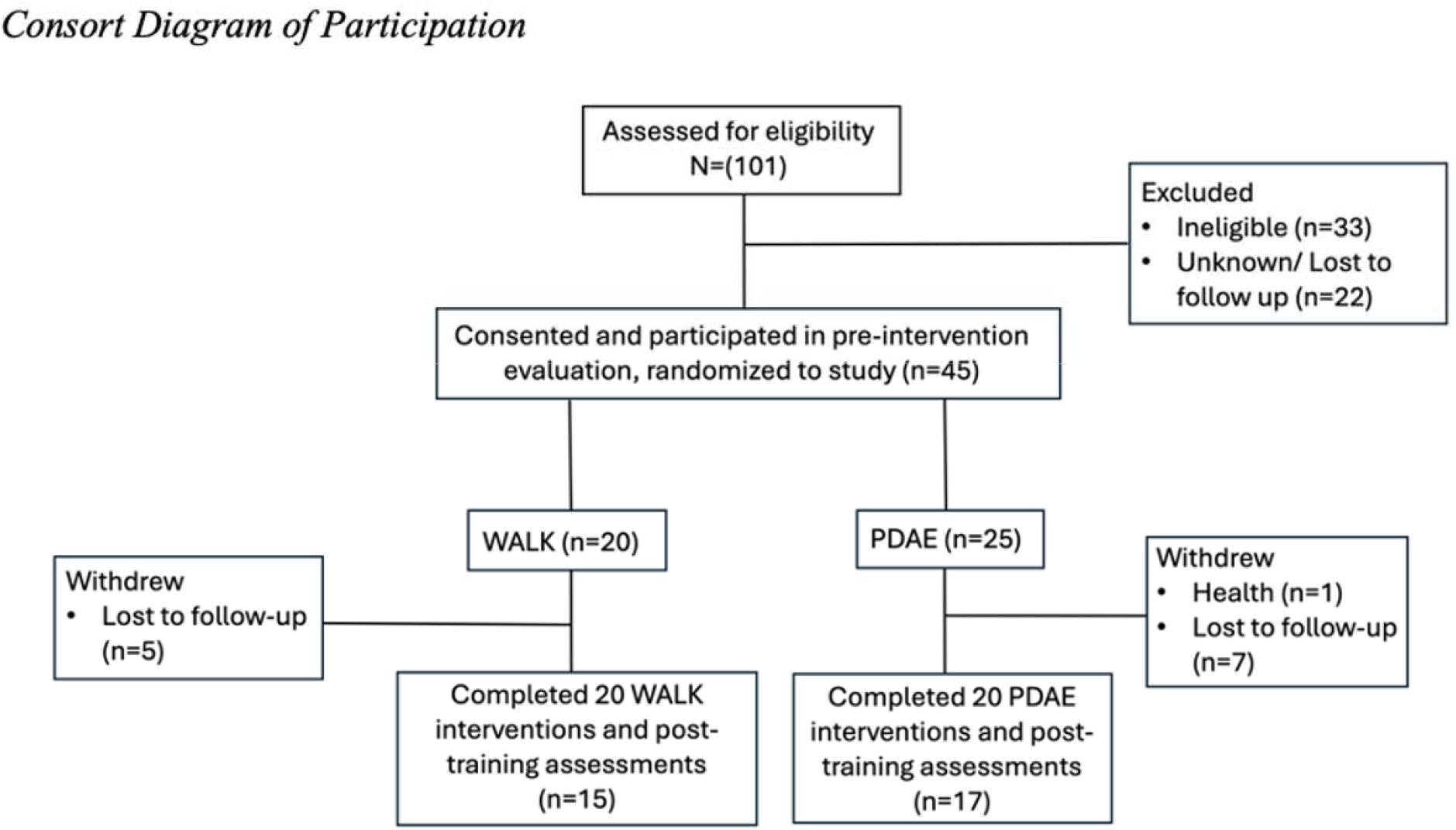
Consort Diagram of Participation.

### Participants

Participants were recruited from the Atlanta VAMC Movement Disorders clinic using the VA Informatics and Computing Infrastructure (VINCI) databases to identify patients in the Atlanta VA Health Care System eligible for the study. Participants were also recruited from the Emory University Movement Disorders clinic, at local Parkinson-related support groups, educational meetings, and community events. Recruitment took place between November 2020 and March 2024. All participants provided written informed consent.

Participants met the following inclusion criteria: 1) a diagnosis of PD (ICD-10 G20) based on established criteria and determined by a board-certified neurologist with training in movement disorders; 2) asymmetric symptoms including at least three of the cardinal signs of PD (rigidity, bradykinesia, tremor, postural instability) and showed clear symptomatic benefit from anti-parkinsonian medications; 3) aged 40 years or older; 4) a Hoehn & Yahr stage I–III; and 5) reported symptoms during their “off” time indicated by score ≥1 on UPDRS-IV item 4.3 (i.e., time spent in the OFF-state). Exclusion criteria included scoring <17 on the Montreal Cognitive Assessment or ≥18 on the Beck Depression Inventory-II or having another neurodegenerative disease. At initial assessment, participants were evaluated for general health, ability to perform ADLs, fall risk, age, and education. The Montreal Cognitive Assessment (MoCA) was administered to determine cognitive status. Participants were excluded from the study if the MoCA score was less than 17.

Using the REDCap randomization module, stratified by age and sex staff assigned participants randomly to PDAE or WALK after baseline assessments. Investigators and administrative staff, but not instructors, were blinded to treatment assignments. Participants knew their treatment but not whether they were allocated to an experimental or control group.

### Interventions

Fidelity was monitored by the investigators via weekly reports from interventionists and monthly class visits of both investigators. The dose for intervention was measured by the time spent in the class, e.g., one class is a dose of 90 minutes. The volume of the intervention is defined as frequency times duration, and therefore the volume per week was approximately 180 minutes/week for the Training Phase, which is the phase being considered in this work. The intensity of the exercise for PDAE and WALK participants was measured with heart rate monitors.

Intervention dose determinants: In the 3-month Training phase, participants were assigned to 12 weeks of 20 biweekly (90-minute) lessons. Fall incidence has been extremely rare over the past ten years of implementing similar type intervention in PD, but fall prevention was a critical concern. All assistants for both PDAE and WALK were trained in PD-related posture problems, monitoring balance, and anticipating and preventing falls with a 2-hour experiential workshop.

### Partnered Dance-Aerobic Exercise (PDAE)

During PDAE, people with PD partnered with an individual without PD, e.g., a trained caregiver, friend, or university student. The instructor and several trained assistants carefully monitored all participants. Participants danced both the follower and leader roles alternatingly, as has been done for several prior studies [11]. They also danced with new partners every 15 minutes; a widely practiced method considered by the dance teaching community to enhance learning. Participants engaged in partnering exercises concerning how to interpret motor goals through touch, exercises to develop understanding temporal relationship of movement to music, novel step introduction, connecting previously learned and novel step elements. Participants were not required to memorize specific step patterns but learned new steps in each class.

### Walking Aerobic Exercise (WALK)

Participants in WALK received the equivalent dose, volume, frequency, intensity, and duration of exercise to the PDAE group. Participants received equal contact and monitoring from study staff. WALK participants reported to the same facility and interacted with the same interventionist and assistants. Participants began the classes with a 25-minute warm-up followed by a 45-minute walking session and a 10-minute cool down, with breaks in between for transitions. The WALK is well described in Wells et al., 2023 [12]. Participants walked outside primarily overground, except when it rained or weather was otherwise inclement, and then participants would walk inside down long hallways at the center where the studies took place. WALK also took place in groups, with research volunteers and assistants, to ensure that PDAE and WALK participants received a socially engaging intervention.

### Evaluations

#### Disease Severity and OFF-time

A blinded MDS-certified rater administered the MDS-UPDRS-I-IV at all evaluations. The MDS-UPDRS has 4 parts. I: non-motor experiences of daily living, II: motor experiences of daily living, III: motor examination, and IV: motor complications. The primary outcome, the MDS-UPDRS-IV score, measures MRMF, including dyskinesia, OFF-time, functional impact and complexity of fluctuations, and dystonia. MDS-UPDRS-IV item 3 measures the time spent in the OFF state, and item 4 measures the functional impact of the motor fluctuations. Each question is scored on a 5-point scale: (0) normal, (1) slight, (2) mild, (3) moderate, and (4) severe. A higher total score corresponds to more severe impairment. We administered a monthly 3-day OFF-state diary for corroboration. OFF-state diaries were sent out via email to participants at the beginning of each month, and reminders texts and calls were sent out each week to ensure completion. Participants were first educated about the different states (asleep, ON, ON with troubling dyskinesia, or OFF) and then filled out the diary half-hourly over three consecutive days. Percent waking hours spent in each state were calculated. Percent waking hours spent in the OFF-state was determined as percent waking hours spent in the OFF-state plus percent waking hours spent in the ON with troubling dyskinesia state.

#### Self-Administered Psychosocial Questionnaires – QOL and Independence Measures

To measure the psychosocial impact of the social support/human touch interactions inherent to partnered dance exercise, participants completed several questionnaires within the week before each visit to complete questionnaires i.e., Beck Depression inventory–II (BDI-II), the Physical Activity Scale for the Elderly (PASE), the PD Questionnaire-39 (PDQ-39), the Short Form 12 Health Survey (SF-12), Freezing of Gait questionnaire (FOGQ), and Gait and Falls questionnaire.

### Statistical Analysis

Descriptive statistics for all demographic, clinical, and outcome measures were calculated and reviewed visually. Because all data was normally distributed, independent t-tests were conducted to determine the change in outcome performance overtime (post-training minus baseline) between groups (PDAE and WALK). Paired t-tests were conducted to determine the change in outcome performance within groups (PDAE or WALK) between timepoints (baseline to post-training). A Pearson correlation coefficient was computed to assess the linear relationship between the percent change in percentage of waking hours spent in OFF-state and QOL and independence measures. These analyses were performed using STATA version 18 (StataCorp, College Station, Texas, USA). Because this was a longitudinal study where repeated measures were taken on the same subject, each outcome variable was further analyzed with a separate linear mixed model with a fixed effect for Time and Group, an interaction between Time and Group, and a random effect for participant. Linear mixed models were implemented in lmertest::lmer in RStudio version 2023.06.0 (RStudio, Boston, Massachusetts, USA).

## Results

Forty-five participants (73.33% male) were randomized (PDAE, n = 25; WALK, n = 20). Of the initial randomized participants, thirty-two completed the study through post-training (PDAE, n = 17; WALK, n =15). Descriptive and demographic statistics of the sample are summarized in Table 1. Because of the randomization, our cohorts were relatively similar at baseline.

**Table 1.**
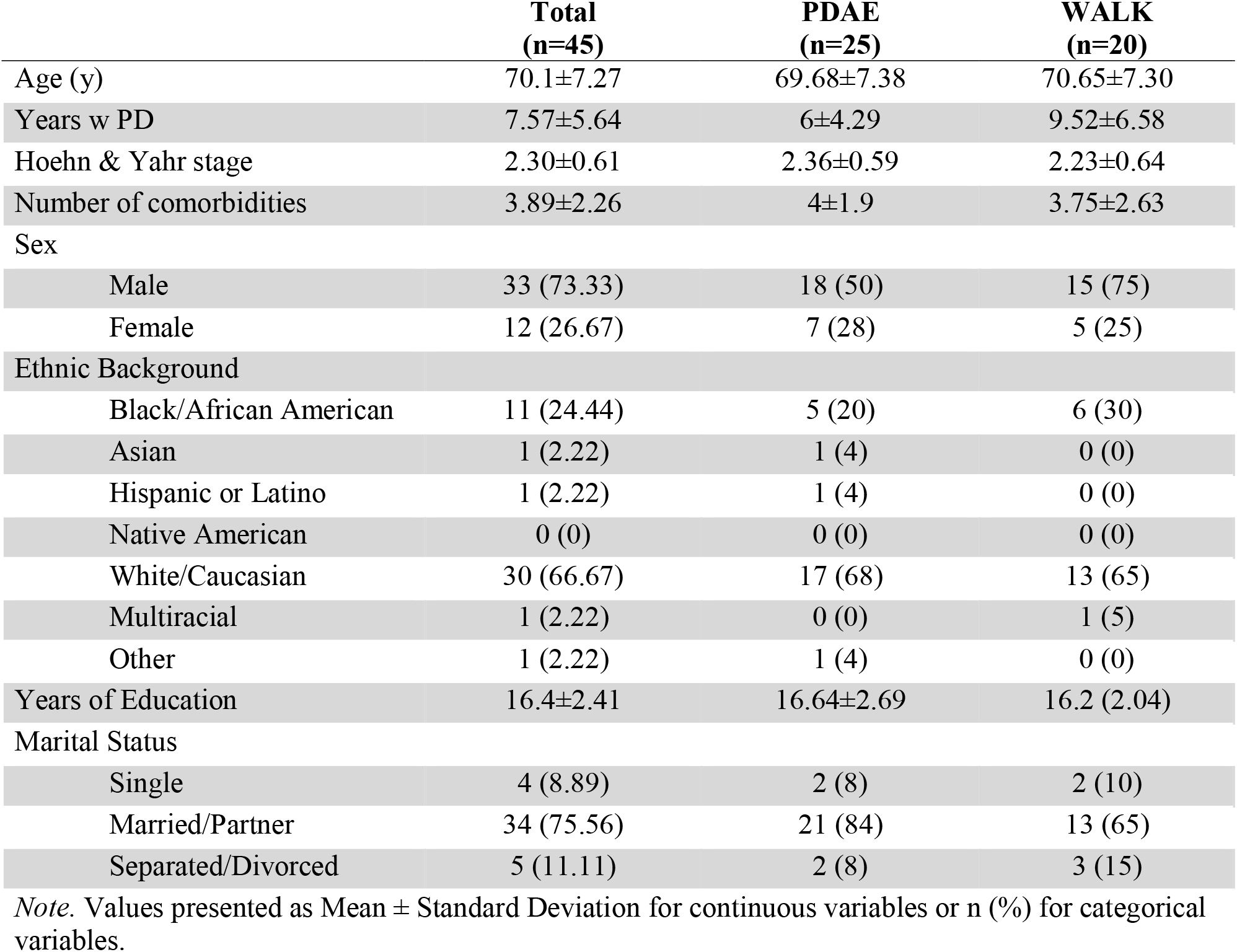
Participant’s Demographic and Clinical Characteristics at Baseline.

### Baseline to post-training

In the PDAE group, there was a significant change in MDS-UPDRS-III (*p* = 0.02). The PDAE group also had a significant change in MDS-UPDRS-IV item 3 (*p* = 0.02), MDS-UPDRS Part IV item 4 score (*p* = 0.02), and MDS-UPDRS Part IV total score (*p* = 0.02, Table 3). No changes were noted in the QOL and independence measures in either group (Table 2). No changes were noted in the WALK group.

**Table 2.**
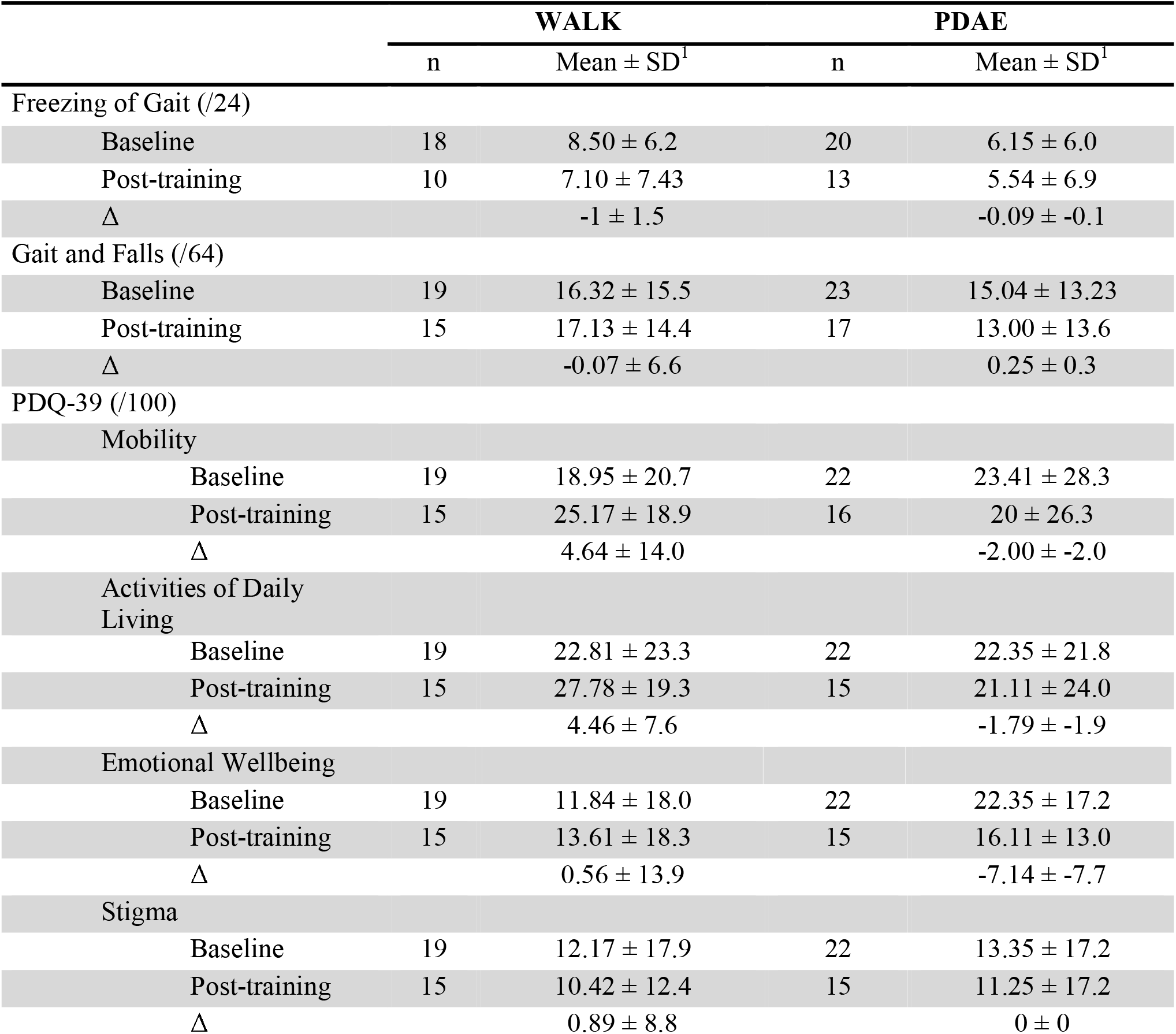
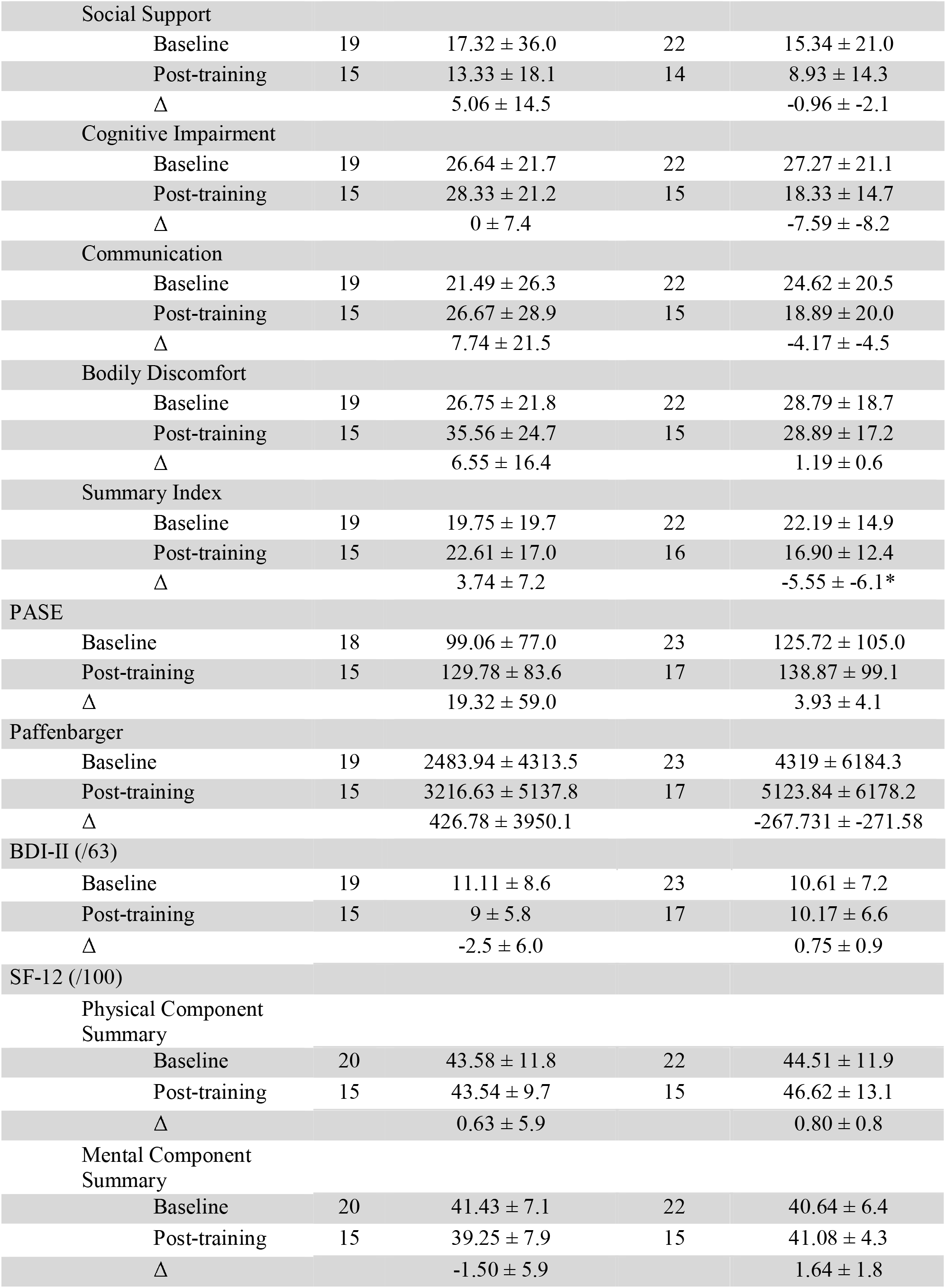
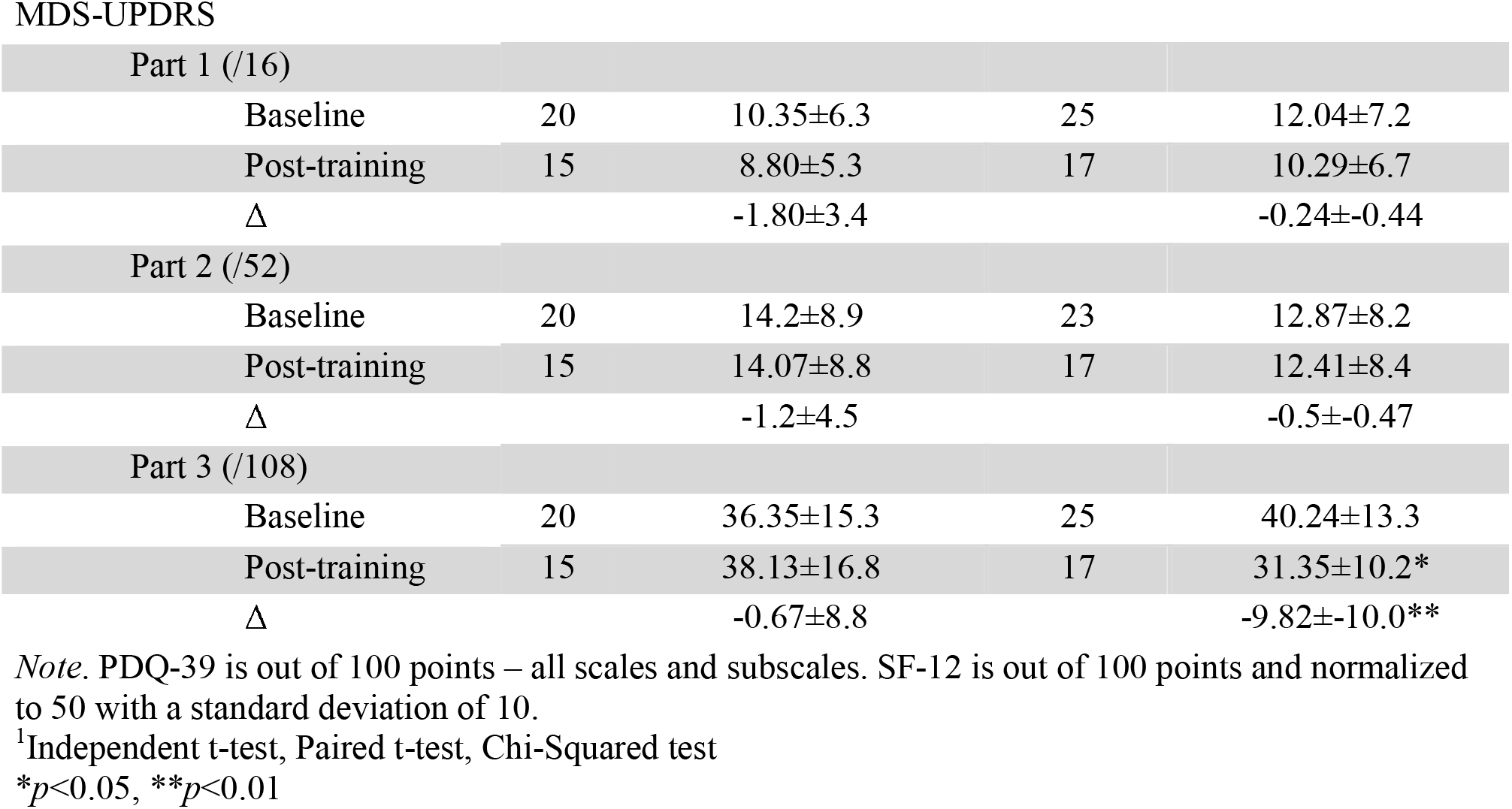
WALK versus PDAE Baseline, Post-training, and Δ in QOL/Independence Outcome Measures.

|  | WALK |  | PDAE |  |
| --- | --- | --- | --- | --- |
| | n | Mean ± SD<sup>1</sup> | n | Mean ± SD<sup>1</sup> |
| Freezing of Gait (/24) |  |  |  |  |
| Baseline | 18 | 8.50 ± 6.2 | 20 | 6.15 ± 6.0 |
| Post-training | 10 | 7.10 ± 7.43 | 13 | 5.54 ± 6.9 |
| $\Delta$ | | -1 ± 1.5 | | -0.09 ± -0.1 |
| Gait and Falls (/64) |  |  |  |  |
| Baseline | 19 | 16.32 ± 15.5 | 23 | 15.04 ± 13.23 |
| Post-training | 15 | 17.13 ± 14.4 | 17 | 13.00 ± 13.6 |
| $\Delta$ | | -0.07 ± 6.6 | | 0.25 ± 0.3 |
| PDQ-39 (/100) |  |  |  |  |
| Mobility |  |  |  |  |
| Baseline | 19 | 18.95 ± 20.7 | 22 | 23.41 ± 28.3 |
| Post-training | 15 | 25.17 ± 18.9 | 16 | 20 ± 26.3 |
| $\Delta$ | | 4.64 ± 14.0 | | -2.00 ± -2.0 |
| Activities of Daily Living |  |  |  |  |
| Baseline | 19 | 22.81 ± 23.3 | 22 | 22.35 ± 21.8 |
| Post-training | 15 | 27.78 ± 19.3 | 15 | 21.11 ± 24.0 |
| $\Delta$ | | 4.46 ± 7.6 | | -1.79 ± -1.9 |
| Emotional Wellbeing |  |  |  |  |
| Baseline | 19 | 11.84 ± 18.0 | 22 | 22.35 ± 17.2 |
| Post-training | 15 | 13.61 ± 18.3 | 15 | 16.11 ± 13.0 |
| $\Delta$ | | 0.56 ± 13.9 | | -7.14 ± -7.7 |
| Stigma |  |  |  |  |
| Baseline | 19 | 12.17 ± 17.9 | 22 | 13.35 ± 17.2 |
| Post-training | 15 | 10.42 ± 12.4 | 15 | 11.25 ± 17.2 |
| $\Delta$ | | 0.89 ± 8.8 | | 0 ± 0 |
| Social Support |  |  |  |  |
| Baseline | 19 | 17.32 ± 36.0 | 22 | 15.34 ± 21.0 |
| Post-training | 15 | 13.33 ± 18.1 | 14 | 8.93 ± 14.3 |
| Δ |  | 5.06 ± 14.5 |  | -0.96 ± -2.1 |
| Cognitive Impairment |  |  |  |  |
| Baseline | 19 | 26.64 ± 21.7 | 22 | 27.27 ± 21.1 |
| Post-training | 15 | 28.33 ± 21.2 | 15 | 18.33 ± 14.7 |
| Δ |  | 0 ± 7.4 |  | -7.59 ± -8.2 |
| Communication |  |  |  |  |
| Baseline | 19 | 21.49 ± 26.3 | 22 | 24.62 ± 20.5 |
| Post-training | 15 | 26.67 ± 28.9 | 15 | 18.89 ± 20.0 |
| Δ |  | 7.74 ± 21.5 |  | -4.17 ± -4.5 |
| Bodily Discomfort |  |  |  |  |
| Baseline | 19 | 26.75 ± 21.8 | 22 | 28.79 ± 18.7 |
| Post-training | 15 | 35.56 ± 24.7 | 15 | 28.89 ± 17.2 |
| Δ |  | 6.55 ± 16.4 |  | 1.19 ± 0.6 |
| Summary Index |  |  |  |  |
| Baseline | 19 | 19.75 ± 19.7 | 22 | 22.19 ± 14.9 |
| Post-training | 15 | 22.61 ± 17.0 | 16 | 16.90 ± 12.4 |
| Δ |  | 3.74 ± 7.2 |  | -5.55 ± -6.1* |
| PASE |  |  |  |  |
| Baseline | 18 | 99.06 ± 77.0 | 23 | 125.72 ± 105.0 |
| Post-training | 15 | 129.78 ± 83.6 | 17 | 138.87 ± 99.1 |
| Δ |  | 19.32 ± 59.0 |  | 3.93 ± 4.1 |
| Paffenbarger |  |  |  |  |
| Baseline | 19 | 2483.94 ± 4313.5 | 23 | 4319 ± 6184.3 |
| Post-training | 15 | 3216.63 ± 5137.8 | 17 | 5123.84 ± 6178.2 |
| Δ |  | 426.78 ± 3950.1 |  | -267.731 ± -271.58 |
| BDI-II (/63) |  |  |  |  |
| Baseline | 19 | 11.11 ± 8.6 | 23 | 10.61 ± 7.2 |
| Post-training | 15 | 9 ± 5.8 | 17 | 10.17 ± 6.6 |
| Δ |  | -2.5 ± 6.0 |  | 0.75 ± 0.9 |
| SF-12 (/100) |  |  |  |  |
| Physical Component Summary |  |  |  |  |
| Baseline | 20 | 43.58 ± 11.8 | 22 | 44.51 ± 11.9 |
| Post-training | 15 | 43.54 ± 9.7 | 15 | 46.62 ± 13.1 |
| Δ |  | 0.63 ± 5.9 |  | 0.80 ± 0.8 |
| Mental Component Summary |  |  |  |  |
| Baseline | 20 | 41.43 ± 7.1 | 22 | 40.64 ± 6.4 |
| Post-training | 15 | 39.25 ± 7.9 | 15 | 41.08 ± 4.3 |
| Δ |  | -1.50 ± 5.9 |  | 1.64 ± 1.8 |

MDS-UPDRS
| Part 1 (/16) |  |  |  |  |
| --- | --- | --- | --- | --- |
| Baseline | 20 | 10.35±6.3 | 25 | 12.04±7.2 |
| Post-training | 15 | 8.80±5.3 | 17 | 10.29±6.7 |
| Δ |  | -1.80±3.4 |  | -0.24±-0.44 |
| Part 2 (/52) |  |  |  |  |
| Baseline | 20 | 14.2±8.9 | 23 | 12.87±8.2 |
| Post-training | 15 | 14.07±8.8 | 17 | 12.41±8.4 |
| Δ |  | -1.2±4.5 |  | -0.5±-0.47 |
| Part 3 (/108) |  |  |  |  |
| Baseline | 20 | 36.35±15.3 | 25 | 40.24±13.3 |
| Post-training | 15 | 38.13±16.8 | 17 | 31.35±10.2* |
| Δ |  | -0.67±8.8 |  | -9.82±-10.0** |

*Note.* PDQ-39 is out of 100 points – all scales and subscales. SF-12 is out of 100 points and normalized to 50 with a standard deviation of 10.
<sup>1</sup>Independent t-test, Paired t-test, Chi-Squared test
\* $p < 0.05$ , \*\* $p < 0.01$

### PDAE versus WALK

Linear mixed-effects model revealed that the interaction between group and time had a significant effect on the change in the MDS-UPDRS-III score (*p* = 0.001), MDS-UPDRS-IV item 3 (*p =* 0.02), and MDS-UPDRS-IV total score *(p* = 0.02). The PDAE group had a significant reduction in MDS-UPDRS-III score (p = 0.02), MDS-UPDRS-IV item 3 score (*p* = 0.02), and MDS-UPDRS-IV total score (*p* = 0.03) compared to the WALK group (Table 3).

**Table 3.** WALK versus PDAE Baseline, Post-training, and Δ in OFF-time Outcome Measures.

|  | WALK |  | PDAE |  |
| --- | --- | --- | --- | --- |
|  | n | Mean ± SD<sup>1</sup> | n | Mean ± SD<sup>1</sup> |
| MDS-UPDRS-IV |  |  |  |  |
| Item 3 |  |  |  |  |
| Baseline | 20 | 1.15 ± 0.8 | 25 | 1.6 ± 1 |
| Post-training | 14 | 1.07 ± 0.7 | 17 | 0.88 ± 0.9* |
| Δ |  | -0.07 ± 0.9 |  | -0.82 ± -0.9* |
| Item 4 |  |  |  |  |
| Baseline | 20 | 1.4 ± 1.5 | 25 | 1.48 ± 1.2 |
| Post-training | 14 | 1.00 ± 1.2 | 17 | 0.65 ± 1.0* |
| Δ |  | -0.43 ± 1.3 |  | -0.71 ± 0.8 |
| Total score (/23) |  |  |  |  |
| Baseline | 20 | 6 ± 4.4 | 25 | 6.48 ± 3.7 |
| Post-training | 14 | 5.71 ± 3.15 | 17 | 3.59 ± 3.5* |
| Δ |  | -0.12 ± 2.8 |  | -2.47 ± -2.8* |
| 3-Day OFF-state Diary |  |  |  |  |
| % ON |  |  |  |  |
| Baseline | 14 | 70.45 ± 27.4 | 13 | 71.25 ± 27.3 |
| Post-training | 12 | 78.62 ± 24.2 | 9 | 58.17 ± 33.1 |
| Δ |  | 0.84 ± 17.9 |  | -21.89 ± -24.6 |
| % OFF |  |  |  |  |
| Baseline | 14 | 22.98 ± 19.5 | 13 | 26.26 ± 23.3 |
| Post-training | 12 | 21.06 ± 23.8 | 9 | 41.44 ± 22.1 |
| Δ |  | -0.69 ± 18.3 |  | -2.20 ± -2.5 |

<sup>1</sup>Independent t-test, Paired t-test, Chi-squared test
\* $p < 0.05$

Linear mixed-effects model revealed that the interaction between group and time had a significant effect on the change in PDQ-39 summary index score (*p =* 0.02). The PDAE group also had a significant reduction in PDQ-39 summary index score compared to WALK group (*p* = 0.02, Table 2).

Fixed effects of group, time, and interaction between group and time did not significantly impact the change of all other QOL, independence (Table 2), and OFF-time measures (Table 3) between groups. There was no significant change in scores between groups for the BDI-II survey, PDQ-39 subscales, PASE survey, Paffenbarger survey, SF-12 survey, FOG questionnaire, or Gait and Falls questionnaire. There was no significant change in the percentage of waking hours spent in the ON or OFF-state as reported by the 3-day OFF-state diary or MDS-UPDRS-IV item 4 score between the WALK and PDAE groups.

### Pearson correlation between percent change of waking hours spend int the OFF-state against percent change in all other QOL and independence variables

Pearson correlation (r) and p values between the overall percent change of waking hours spent in the OFF state against the percent change in all other QOL and independence variables are summarized in Table 4. There was a strong correlation with the PDQ-39 social support subscale (r = 0.90, *p*=0.005), suggesting that those who experienced a decrease in time spent in the OFF-state also demonstrated a decreased need for social support in their everyday lives. There was a moderate correlation with percent change in the Paffenbarger survey (r=0.56, *p*=0.03), suggesting that those who experienced a decrease in time spent in the OFF state also demonstrated a decrease in physical activity.

**Table 4.** Pearson Correlations between the percent change of the percentage of waking hours spent in the OFF-state and percent change in QOL and independence measures for both groups.

| Variable | $r^1$ | $p^1$ |
| --- | --- | --- |
| PASE | -0.39 | 0.29 |
| Paffenbarger | 0.56* | 0.03 |
| PDQ-39 |  |  |
| Mobility | -0.23 | 0.47 |
| ADL | 0.14 | 0.14 |
| Emotional Wellbeing | 0.19 | 0.19 |
| Stigma | 0.14 | 0.14 |
| Social Support | 0.90** | 0.005 |
| Cognitive Impairment | -0.01 | 0.97 |
| Bodily Discomfort | -0.17 | 0.55 |
| Summary Index | 0.05 | 0.85 |
| BDI-II | 0.07 | 0.81 |
| SF-12 |  |  |
| Physical Component Summary | 0.09 | 0.72 |
| Mental Component Summary | 0.09 | 0.73 |

<sup>1</sup>Pearson correlation

## Discussion

This study demonstrated improved motor function and OFF-time among 17 individuals with PD who completed 30 hours of adapted Argentine tango in comparison to 15 individuals with PD who completed WALK training for the same duration. With the noted within-group changes, this study demonstrated improved burden of PD to QOL from the PDQ-39 survey, motor function from the MDS-UPDRS-III, and motor complications from the MDS-UPDRS-IV in the PDAE group compared to the WALK group. Overall, a decrease in OFF-time was correlated to a decreased need for social support. Interestingly, a decrease in OFF-time was correlated with less physical activity through the Paffenbarger survey. Whether this correlation is directly attributable to the changes seen after a short program of PDAE or WALK or a third factor is unclear and warrants further investigation. The other findings generally support that PDAE is a feasible and effective form of aerobic exercise for individuals with PD to serve as an adjunctive therapy to current pharmacological methods to improve OFF-time symptoms and motor function and consequently impact their perceived QOL and independence. Walking aerobic exercise, while inferior to PDAE, may also provide benefits; however, a longer and more robust study is necessary to determine the potential impact of walking aerobic exercise on individuals with PD.

Preventing advancements in the Hoehn and Yahr stage and increases in MDS-UPDRS scores is an essential part of maintaining the QOL and independence of individuals with PD. PDAE’s improvement on the MDS-UPDRS-III and IV may suggest that PDAE helps slow disease progression, reduce the severity of motor impairment, and help the experience of motor complications. Typically, functional independence decreases throughout the disease progression. Independent life becomes complicated for people with PD in Stages III and IV (severe disability, unable to walk and stand unassisted) and beyond [13]. Higher MDS-UPDRS scores are associated with worse QOL, specifically higher PDQ-39 summary index scores, mobility, ADL, and communication subscales, the PDQ-8 (short version of PDQ-39) and EuroQOL-5D (five-item generic health status measure) [14].

Because PDAE helped reduce the severity of motor complications, it is unsurprising that the PDQ-39 summary index score also improved through PDAE. Studies have correlated OFF-time and QOL through the PDQ-39 survey. Patients who experienced OFF-time had higher scores on most PDQ-39 subscales and the overall summary index, indicated worse QOL, compared to those who did not experience OFF-time. PDQ-39 scores were also directly related to the number of hours spent in the OFF-state [7]. Our study suggests that improving OFF-time may help enhance the individual’s perception of their QOL by improving all aspects of the PDQ-39 overall, not just specific subscales. However, further studies are needed to better associate the relationship between improving OFF-time and its effects on QOL after PDAE and WALK.

Neither significant improvements nor declines were noted in other QOL or independence measures. As such, the program did lead to maintenance of these measures, which typically worsen due to the neurodegenerative nature of PD. Further, longer studies are needed to truly understand the impact of improving OFF-time on QOL and independence in individuals with PD. In older adults, more consistent findings of structural changes (increase in white and gray matter and volumetric increases) in the brain are seen after at least 6 months of consistent aerobic exercise [15]. Although benefits of exercise on QOL can be seen after three months, typically, studies on PD look at changes in QOL and structural and functional changes in the brain after six to twelve months [16]. Greater QOL, specifically physical health QOL, as it relates to self-reported activities of daily living and self-perceived pain and discomfort, are associated with greater brain integrity in older adults [17].

Adapted tango incorporates several aspects of movement that are possibly relevant for individuals with PD and may explain its superiority to walking aerobic exercise. Tango requires the participants to multitask and requires dynamic balance, turning, initiation of movement, moving at a variety of speeds, and often backward while close to a partner [11]. Using auditory cues, like in PDAE, to help facilitate movement has been proven to be beneficial for individuals with PD. Auditory cues can help increase gait initiation, walking speed, and cadence in laboratory settings, while performing functional tasks at home, and has been shown to reduce the severity of freezing. Auditory cues are particularly beneficial for individuals with PD since they may be able to bypass the defective look from the basal ganglia to the supplementary motor area via the thalamus that is normally used for internally cued movements [18]. Therefore, the use of rhythmic cues from music, and guidance from their partners may be an important feature of tango as an effective intervention for individuals with PD.

Prior work has demonstrated adapted tango’s improvement on mobility and QOL in PD. Along with improving motor symptoms in individuals with PD, dance, in general, may help improve QOL because it reduces social isolation and increases participation in life situations. After a yearlong program of tango, participants recovered lost activities, began new ones, and gained the ability to engage in more complex activities, all of which enhanced their QOL [19].

Additionally, exercise has been found to influence the pharmacokinetics of levodopa. A mice model study found that exercise can partially prevent the development of levodopa-induced dyskinesia. It also attenuates the side effects of levodopa through normalization of the striatopallidal dopaminergic signaling without affecting the anti-parkinsonian effects of levodopa [19]. Exercise can also improve dopamine efficiency. A study found that positron emission tomography (PET) scans of a cohort of naïve PD patients showed that within one year of diagnosis and prior to medication use showed that exercise increased expression of D2 receptors [20]. Therefore, exercise, specifically dance because of its unique factors, might influence the experience of OFF-time by impacting levodopa efficiency.

Although PDAE and WALK both require balance and attention to movement control, tango differs from walking aerobic exercise because it is performed in close relationship to a partner and in a setting that fosters community involvement, it is progressive in nature and the participant is always learning, and it is performed to music which may further engage the participant in addition to serving as an auditory cue [11]. Thus, adapted tango elements, including light-moderate intensity, and structured motor components that engage a memory of steps and directions while encouraging a keen awareness of spatial relationship and rhythm could contribute to some currently undetermined mechanisms to improvements in OFF-time. Further study into the application of therapies such as adapted tango for improving OFF-time is warranted and needed, given the prevalence of OFF-time in PD, which affects QOL and independence.

### Limitations and future directions

Although this study was the first of its kind to demonstrate improved OFF-time measures in addition to the experience of daily living and motor function in individuals with PD who participated in a 3-month program of adapted tango and compared to individuals with PD who participated in walking aerobic exercise, it is not without its limitations. Due to the relatively small sample size and notable attrition rates (32% for PDAE, 25% for WALK), many of the whole study correlations between QOL and independence outcome measures and OFF-time showed no relationship and were not significant, and we are unable to determine if there is an association or correlation within each group. Additionally, since this study is only the first three months of a longer 16-month study, we are unable to determine how the change in OFF-time can predict the change in QOL and independence outcome measures. However, this study is a preliminary report about the first three months of a longer 16-months study. Larger studies are necessary to make more definitive conclusions. Future studies looking at the effect of the full program of 16-months of PDAE versus WALK on individuals with PD will be able to overcome these limitations.

## Conclusion

OFF-time is one of the most disconcerting features of PD and severely impacts the individual’s QOL and independence. This novel study demonstrates that a short program of adapted tango improves motor function, OFF-time, and the experience of daily living in older adults with PD compared to a short program of walking aerobic exercise. This work may ultimately lead to further implementation of therapeutic movement and dance therapy as an effective and enjoyable non-pharmacological strategy for addressing OFF-time in individuals with PD to help improve their QOL and independence.

## Data Availability

All data produced in the present study are available upon reasonable request to the authors

## Funding

A Department of VA Merit Award, 1 I01 RX002967 supports the PAIRED Trial. Dr. Hackney was supported by grant funding from the NIH (NIH-NINDS 1K23NS105944-01A1) and the American Parkinson’s Disease Association Center for Advanced Research at Emory University.

## Competing interests

The authors reported no conflict of interest.

## Data Availability Statement

The data supporting this study’s findings are available from the corresponding author upon reasonable request.

## Notes

### Competing Interest Statement

The authors have declared no competing interest.

### Clinical Trial

NCT04122690

### Funding Statement

This study was funded by the United States Department of VA Merit Award 1 I01 RX002967. Daniel Huddleston was supported by grant funding from the NIH (NIH-NINDS 1K23NS105944-01A1) and the American Parkinsons Disease Association Center for Advanced Research at Emory University.

### Author Declarations

The Institutional Review Board (IRB) of Emory University and the Research and Development Committee of the Atlanta Veterans Affairs Medical Center gave ethical approval for this work.

